# “Professional Identity at the Crossroads: Italian Healthcare Technologists navigating Technology and Human Care”

**DOI:** 10.1101/2025.07.09.25331195

**Authors:** I. Salvatori, L. Mortari, R. Silva

## Abstract

Healthcare Technologists (HTs) represent a crucial yet understudied group within modern healthcare systems. Operating at the intersection of advanced technologies and patient care, HTs face the dual challenge of mastering complex technical procedures while sustaining ethically grounded, relational engagement. This study investigates how professional identity is constructed within HT education in Italy, combining conceptual analysis and empirical data. Drawing on the philosophy of technology and theories of professional identity, the research critiques the traditional technocentric orientation of HT training and advocates for pedagogical models that integrate emotional, ethical, and reflective dimensions.

The empirical component includes 55 semi-structured interviews with 40 HT educators (clinical tutors and adjunct lecturers) and 15 undergraduate students from various HTs academic programs. Using inductive content analysis, the study identifies four macro-categories shaping professional identity: career motivation and background; identity-building processes; relational and ethical values; and educational strategies. Participants describe identity formation as a dynamic, reflective process shaped by real-world practice, relational interactions, and motivational drivers. Both students and educators emphasize the hybrid nature of their role, blending technical mastery with interpersonal care. Relational competence, adaptability, and a commitment to quality emerge as defining features.

The study highlights systemic challenges, including limited recognition of HTs’ roles, pedagogical challenges for educators, and documents strong intrinsic motivations and ethical commitment among participants. HT educators express the need for greater support in transitioning from clinical to teaching roles, and students reveal a desire for more hands-on and reflective learning experiences.

This research contributes a conceptual and practical framework for rethinking HTs education, embedding humanistic, ethical, and relational competencies into curricula. It positions HTs not as peripheral technicians, but as integral professionals in technologically mediated, patient-centered care systems.

## Introduction

Healthcare Technologists (HTs) are a vital and often underexamined component of modern healthcare systems. These professionals operate at the intersection of advanced diagnostic and therapeutic technologies and direct patient care. While their technical expertise is essential, their roles also demand significant relational, communicative, and ethical engagement, particularly in emotionally charged clinical contexts such as oncology, intensive care, pediatrics, and geriatrics.

Their professional identity is shaped by this dual responsibility: delivering high-precision technical services while remaining attuned to the human dimension of healthcare. Despite this hybrid nature, HTs remain largely absent from theoretical models and empirical studies in Health Professions Education (HPE), which have traditionally focused on physicians and nurses. These professions are supported by pedagogical frameworks that emphasize reflective practice, ethical reasoning, and interpersonal communication (1,3). In contrast, HTs education has often been shaped by a technocentric orientation that privileges procedural mastery and operational efficiency.

In Italy, academic training for HTs is formally defined by Ministerial Decrees n. 270/2004 ^2^, which regulate the Italian Class L/SNT3, Technical Health Professions (Diagnostic and Care). This class includes the following professional profiles:

- Radiographer or Radiology Technologist
- Magnetic Resonance Imaging Technologist
- Computed Tomography Technologist
- Radiation Therapy Technologist
- Nuclear medicine Technologist
- Sonographer
- Medical Laboratory Technologist
- Neurophysiopathology Technician
- Prosthetist and Orthopaedic Technician
- Audiometrist
- Audioprosthetist
- Cardiovascular Technician and Perfusionist
- Dental Hygienist
- Dietitian

Each of these roles involves distinct yet overlapping technical responsibilities and direct patient interaction. HTs interpret clinical data, operate and monitor sophisticated devices, explain procedures to patients, and often support them emotionally during diagnosis or treatment. Nevertheless, although comprehensive empirical studies are lacking, academic training and educational policies in this field appear to have historically emphasized technical instruction, while the development of ethical sensitivity, communication skills, and reflective capacities may not always have been addressed in a structured or systematic way.

To better frame this issue, we draw on the *philosophy of technology*, an interdisciplinary field that critically explores how technology shapes human perception, action, and moral agency. Philosophers such as Don Ihde and Peter-Paul Verbeek argue that technology is not a passive instrument but an active mediator in human-world relationships (4,5). In the clinical context, this implies that medical devices, artificial intelligence (AI) systems, and diagnostic software not only support clinical reasoning but also condition how professionals perceive patients, engage with care processes, and experience their own role (6). This insight raises important pedagogical questions: How can we prepare HTs to critically reflect on the technological mediation of care? How do we avoid the risk of depersonalization and ethical erosion in increasingly technologized environments?

Recent regulatory changes reinforce the urgency of these questions. In 2025, Italy’s National Federation of Orders for Radiologic Technologists and Other Health Technical Professions (FNO TSRM e PSTRP) approved revised ethical codes ^3^ for 16 healthcare professions, including all L/SNT3 profiles, and the ethical constitution^4^. These documents, developed through a participatory process, reflect a renewed commitment to principles such as digital ethics, responsible use of artificial intelligence, gender-sensitive care, safety, and equitable access. The new codes underscore the need for HTs to engage critically with technology and reaffirm the primacy of patient-centered care.

This paper aims to fill a critical gap in HPE by proposing a conceptual framework for understanding HT professional identity as a process of *technological-human integration*. We combine insights from the philosophy of technology with theories of professional identity formation to argue that HT education must move beyond procedural training toward pedagogies that promote emotional resilience, narrative competence, ethical awareness, and reflective practice (7,8).

This study investigates how students and educators in Italy experience the construction of professional identity within HT programs. Through a qualitative analysis of lived educational experiences, we explore how future HTs learn to navigate the tension between technology and human care, and what kinds of pedagogical strategies may best support them in becoming both competent and ethical professionals.

Through qualitative content analysis, we explore how future HTs experience and negotiate the tension between technology and human care. Our findings suggest concrete strategies for rethinking curricula and supporting educators, many of whom transition to teaching from clinical practice without formal pedagogical training. In doing so, we contribute to the broader effort of ensuring that HTs are not merely trained as technicians, but prepared to act as ethically grounded, relationally competent healthcare professionals in a technologically complex landscape.

To prepare HTs for the realities of contemporary healthcare, curricula must include training in communication, emotional resilience, bioethics, narrative competence, and reflective practice. Pedagogical models should evolve to support HT educators also in education or mentoring.

## Methods

Building on the need to understand how Healthcare Technologists (HTs) are supported in becoming ethically grounded and relationally competent professionals, this study explores the processes of professional identity formation within HT education. The research is based on 55 in-depth semi-structured interviews with 40 HTs in educational roles, both academic lecturers and clinical tutors, and 15 undergraduate students enrolled in degree programs of Technical Health Professions.

This study adopts a qualitative methodological framework grounded in phenomenology, with the aim of exploring how individuals experience and make sense of their professional identity formation within HTs education. Phenomenology provides a lens through which to investigate the subjective and intersubjective dimensions of identity development, focusing on the meanings that educators and students attribute to their educational and professional trajectories. By prioritizing participants’ lived experiences, this approach allows for a deeper understanding of the relational, ethical, and contextual factors that shape the emergence of professional identity in both academic and clinical learning environments (9,10).

The interview protocol was designed to provide flexibility while ensuring coverage of key thematic areas (11,12). Educators were invited to reflect on their career paths, teaching responsibilities, and conceptions of professional identity, with particular attention to the values they aim to transmit through education. Students were asked to share their motivations for entering the profession, meaningful moments during clinical placements, and the ways in which they integrate technical and relational skills. Both groups were encouraged to discuss interpersonal approaches in clinical settings, perceived challenges in educational pathways, and suggestions for improving the pedagogical model.

Interviews were conducted between February and April 2025, across multiple academic institutions in Italy, one-on-one, in person. All interviews were audio-recorded with consent, transcribed verbatim, and anonymized.

Data were analysed using inductive content analysis (13, 14). The research team coded the transcripts iteratively, first working independently and then meeting for collaborative discussions to consolidate categories and sub-themes. A comparative coding matrix was developed to distinguish between educator and student perspectives while also identifying areas of overlap. This approach allowed for the development of themes that captured not only conceptual understandings but also the emotional and experiential dimensions of identity formation.

### Participants

The participant pool included 40 Healthcare Technologist (HT) educators and 15 undergraduate students from a wide range of disciplines within the Italian L/SNT3 degree class.

The 40 educators represented several professional specializations, with the majority being Radiographers (50%), followed by Medical Laboratory Technologists (15%), Dieticians (13%), and four other specializations (5% each) (Figure 1).

**Figure 1.**
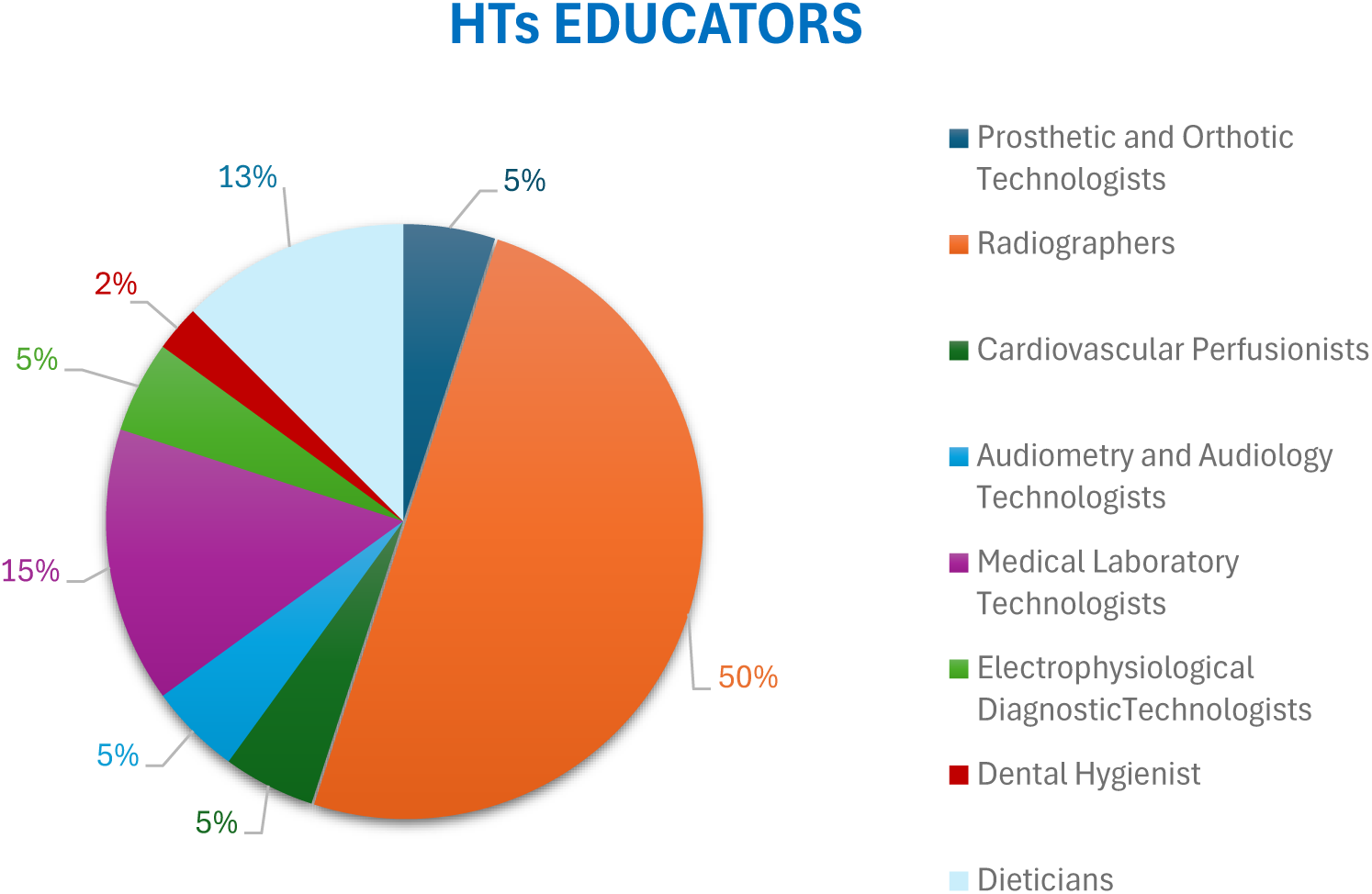
Distribution of educator participants by professional identity.

These educators were evenly divided by institutional role, with 50% serving as Adjunct Healthcare Lecturers and 50% as Clinical Tutors (Figure 2).

**Figure 2.**
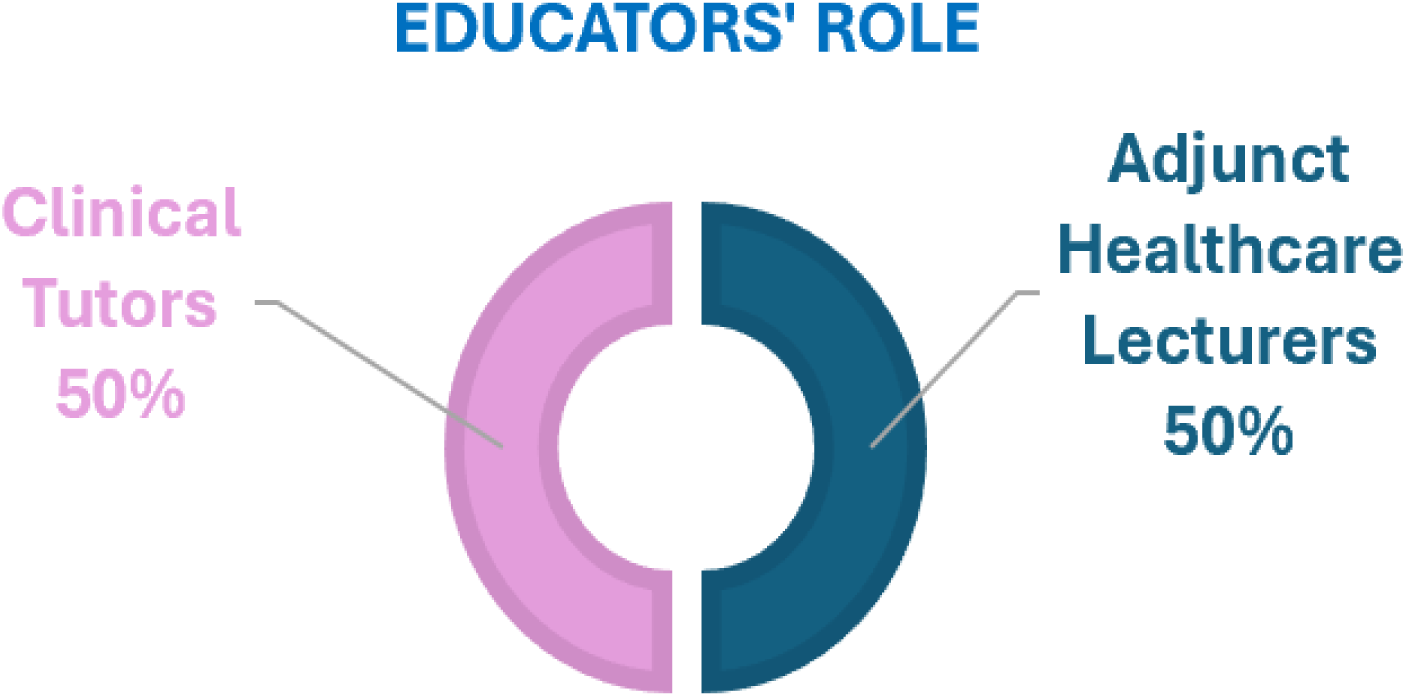
Distribution of educators by role.

These two roles operate under distinct Italian normative frameworks, with Adjunct Lecturers providing formal academic instruction and Clinical Tutors supervising experiential learning in clinical settings.^5^

The 15 students were enrolled in four different educational pathways: Prosthetic and Orthotic Technologists (29%), Radiographers (29%), Electrophysiological Diagnostic Technologists (21%), and Dental Hygienists (21%) (Figure 3).

**Figure 3.**
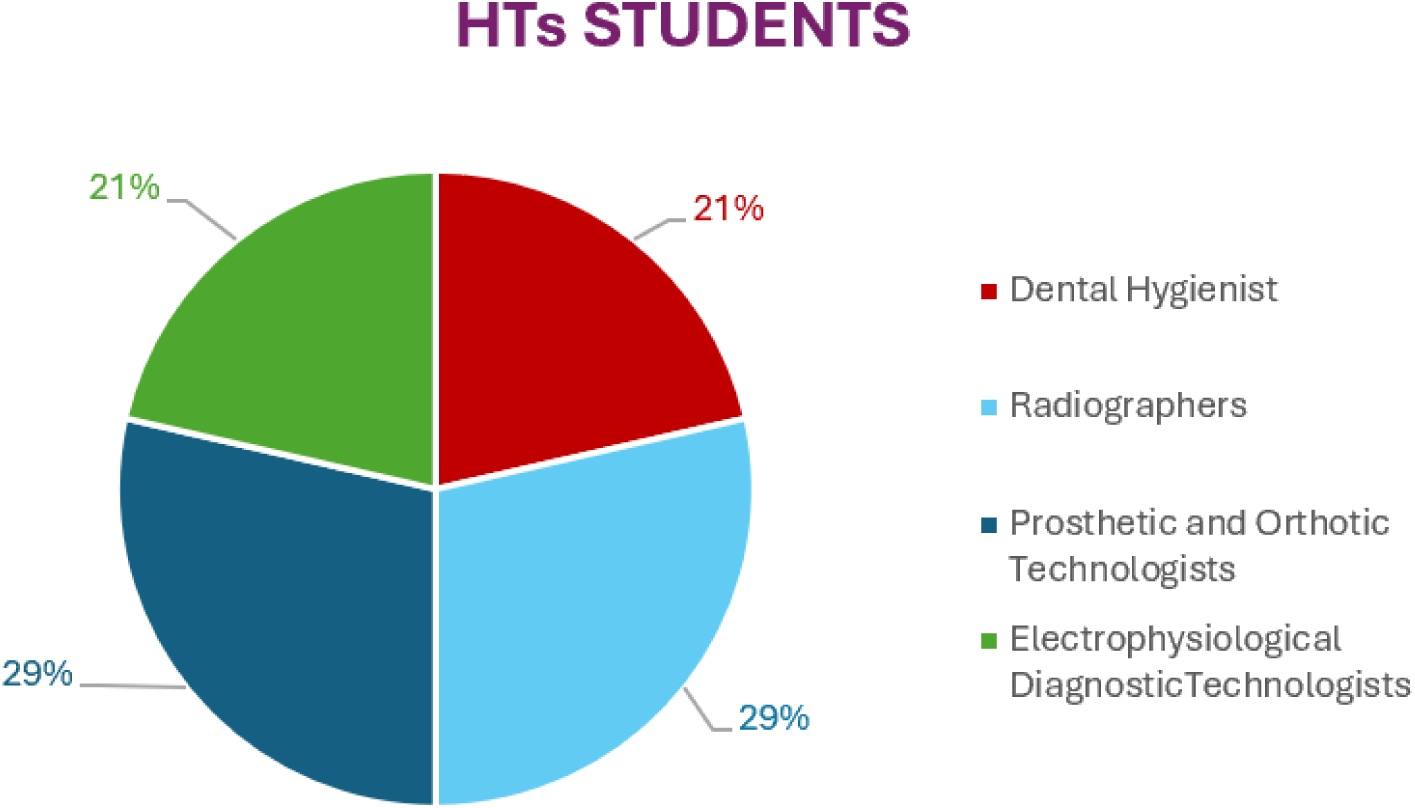
Distribution of student participants by educational pathway.

This disciplinary diversity across both groups was intentional, enabling a comparative exploration of identity formation across various technological and patient-care contexts.

### Recruitment and Ethical Considerations

All participants were recruited through academic and professional networks. Interviews were conducted in Italian, the participants’ native language, to facilitate rich narrative accounts. The study received ethical approval from the Ethics Committee of the Department of Human Sciences at the University of Verona (Record No. 2024_25; Protocol No. EDU-HPTs). All participants provided informed consent, and data were anonymized to ensure confidentiality.

## Results

### Data analysis

The interview data were thematically analyzed and grouped into four macro-categories, reflecting core dimensions of professional identity of HTs. These include:

- **Professional background,** which includes participants’ reasons for choosing the profession, their prior academic or work experiences, and personal narratives linked to their educational journey.
- **Professional identity building**, the most prominent category, which involves growing awareness and formation of one’s professional self, essential role-defining elements (such as the integration of technical and interpersonal skills), aspirations for professional advancement (like enhancing technical skills, teaching abilities), and fundamental professional values (including empathy, ethical practice, and patient focus).
- **Construction of the relational dimension,** which addresses both patient-related interactions (such as fostering a welcoming atmosphere and communicating with clarity and empathy), relationships with students (e.g., fostering student-centred teaching, building rapport and trust) and healthcare team.
- **Foundational elements of educator role and educational path**, referring to the competencies, principles, and approaches necessary to fulfil their respective functions effectively. This includes fostering analytical thinking, commitment to ongoing education, the integration of digital tools in teaching, and active involvement of students.

The resulting coding framework is structured to compare the two participant groups. While separate coding paths were initially developed for educators and students, they are presented together in a unified structure for clarity (see Figure 4 for the coding system diagram).

**Figure 4.**
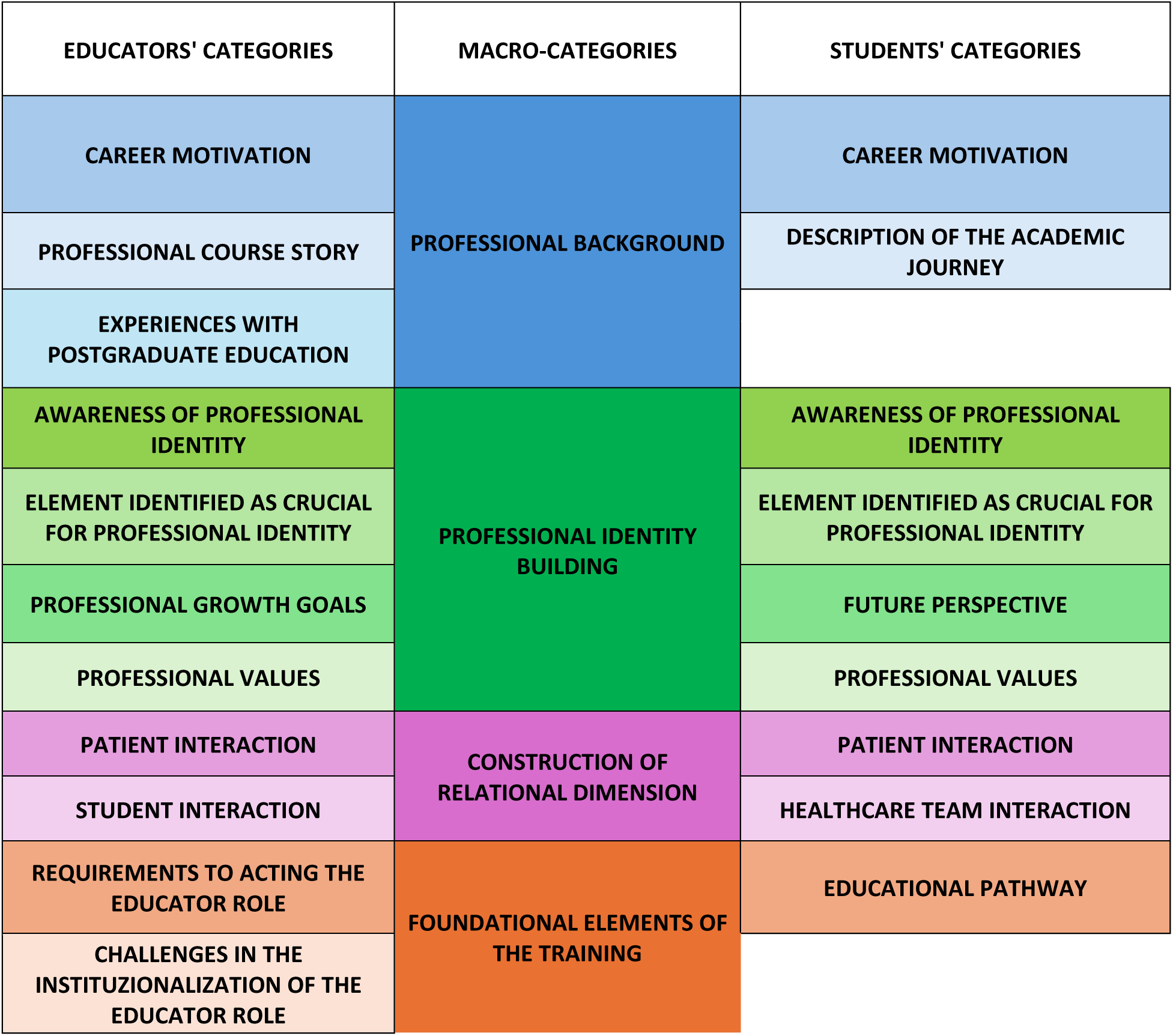
Coding System.

The full descriptive labels for all categories are available in the supplementary materials.

These categories underscore the multidimensional nature of professional identity construction among HTs, revealing how technical expertise is interwoven with relational care, reflective practice, and personal motivation. The structured coding of participants’ responses enabled the study to map the intersections and evolution of these dimensions across various stages of both educational and professional development.

## 1. Professional Background of Educators and Student

### 1.1. Carrer motivation

As evidenced by the interviews, the decision to pursue a career in the healthcare technological fields often stems from a complex interplay between personal inclination and meaningful life experiences. Participants’ career motivations emerge from four distinct pathways: re-evaluating previous choices, pursuing early interests, being inspired by personal encounters, and connecting with profound biographical experiences.

First, for the majority of students (n=8, 53%), the decision to become an HT was the result of a deliberate **re-evaluation of a previous academic or professional path**. These transitions were often driven by a search for more “hands-on and meaningful” work after experiences in other fields proved to be a poor fit. Recalling their moments of realization and how these transitions were often emotionally charged and empowering:

> *“Initially I wanted to be a nurse. I enrolled in the academic course, but after three months of clinical training, I realized it wasn’t for me” (Stud.04.3)*^6^;
>
> *“I started a degree in aerospace engineering, but after Covid and some personal doubts, I realized it wasn’t the right fit. I needed something more hands-on and meaningful” (Stud.06.2);*
>
> *“It was hard to admit pharmacy wasn’t right for me, but I felt anxious during laboratory practice. So, choosing to become a radiographer was the right decision. It aligns better with what I see myself doing” (Stud.02.2)*.

One educator expressed analogous reflections, describing how his vocation for teaching emerged from a need to give back, *“after twenty years in the profession, a desire emerged to leave a mark, a legacy, to share what I had learned in the field” (Ed.04.3)*.

Second, for some participants (n=16, 30%), motivation was rooted in a **long-standing interest in science and the human body**. This early vocation often involved a preference for the “technical and applied” aspects of healthcare over other medical roles. Both students and educators noted:

> *“I’ve always been fascinated by the human body. I considered medicine, but I realized I preferred something more technical and applied” (Stud.07.2);*
>
> *“Science, physics, and anatomy were my favourite subjects. That’s what led me to explore radiography” (Stud.05.03);*
>
> *“I have always been fascinated by the human body; ever since I was a child, I used to read medical encyclopaedias. I had no doubt that I wanted to pursue something in the healthcare field” (Ed.11.04)*.

A recurring theme across participants was an appreciation for the **dual nature of the HT role**, which uniquely combines technical proficiency with direct patient interaction (n=20, 36%). These narratives captured this balance perfectly:

> *I like that this job is so technical and lets me use my hands, work with devices, but also interact with patients” (Stud.05.4);*
>
> *“I’ve always enjoyed working on machineries and device with my hands, but at the same time building relationships with patients. I believe our profession is unique in this regard” (Ed.09.5)*.

Third, **personal encounters with practicing HTs** often served as powerful catalysts (n=12, 21%). Seeing a professional’s passion and understanding their work firsthand was frequently cited as the definitive moment of choice. Some participants were inspired by a family friends:

> *“…a radiology technician, who is my brother’s friend, invited me to spend an afternoon in his clinic, after I finished school. After seeing how he worked, I just said: I want to do this” (Stud.10.05);*
>
> *“My mom’s friend is a dental hygienist. I started asking questions, and seeing her passion really inspired me” (Stud.03.05);*
>
> *“…when I was in high school, I met a lab technician who showed me what his work involved. That’s when I understood what I truly wanted to do” (Ed.09.06)*.

In the last label some narratives, connect past life events and encounters directly to the professional role career motivation:

> *“I was born with agenesis of my left foot, so I’ve always been in contact with orthopaedic services. This degree seemed to unite my personal story with my passion for technical-manual work and the healthcare world. I used to associate orthopaedic technicians with a negative lived experience, but after meeting a very skilled one during my adolescence, I realized the importance of being a quality professional. I saw myself in that role and decided to pursue it” (Stud.13.02);*
>
> *“I come from a family of doctors. But I wanted to build something of my own, in a different field but still close to patients. Teaching came later, as a way of giving back” (Ed.26.02)*.

## 2. Professional Identity Building of Educators and Student

Based on the testimonies reported in the dataset, the macro-category Professional Identity Building clearly stands out as the core of the formative and transformative experience for HTs, both students and educators.

### 2.1. Awareness of one’s professional identity

The development of professional identity emerged as a core transformative experience for both students and educators. Participants consistently described this identity not as a given, but as something earned and constructed over time through four key mechanisms: direct clinical experience, reflective practice and feedback, navigating social perceptions, and overcoming high-stakes challenges.

Among all participants, the **direct clinical experience** was identified as the primary catalyst for identity formation (n=36, 65%). For students, hands-on engagement transformed theoretical knowledge into a tangible professional reality, often solidifying their career choice, *“…but once I started, I realized it was something I truly wanted to do, something I was genuinely passionate about.” (Stud.01.3)*, illustrating how initial uncertainty evolved into intrinsic motivation through practical exposure, *“…the clinical training was very formative; it made me feel part of a team, with all its challenges, mistakes, and unexpected events” (Stud.03.4),and* emphasizing how confronting clinical challenges is central to professional growth.

This process was often emotionally intense but ultimately empowering, marking a clear turning point in their adaptation to the role for both students and educators:

> *“…so once you manage to get the hang of it and understand how the job works, you reach a point where, for example, the clinical training becomes smooth, and now I no longer feel the heaviness of anxiety” (Stud.02.7);*
>
> *“I had met this patient, years earlier when I was volunteering as a paramedic. That morning, while I was on duty, I saw him again, he came in for a nuclear medicine diagnostic exam. Interacting with him was different, […] I not only thought about doing a good diagnostic examination, but also about interpreting the patient’s lived experience” (Ed.03.8);*
>
> *“…it was a great life lesson both as a healthcare professional and as a person […] being able to understand the difficulties faced by the family of this children affected by cerebral palsy” (Ed.01.9)*.

Within this label, it is, in our opinion, important to highlight the role of the reflective practice, often triggered by external feedback, for consolidating this emerging identity from educators (n=13, 32%). This included formal and informal feedback from students and colleagues, which prompted self-assessment. As one educator noted, *“…maybe it’s the feedback you receive from daily work, even from students, that constantly helps you reflect on who you are as a healthcare professional” (Ed.06.8).* Being acknowledged by other professionals was a particularly powerful form of validation: *“…during that surgical procedure, in that tense moment, it was gratifying for me because the physician considered that me, another professional, specifically, a perfusion technologist, could offer their opinion” (Ed.07.6).* Inspirational role models, such as former tutors who later became colleagues, also embodied the professional values that participants sought to internalize and pass on: *“…one of my tutors during university later became my colleague. He embodied what a tutor should be, generous with his time, passionate about sharing both professional knowledge and human values. […] I now feel the responsibility to pass on to students what I was fortunate to receive from her.” (Ed.18.7)*.

A considerable number of participants (n=10, 25%) reported also the challenge of navigating social perceptions and professional boundaries in everyday clinical experience, in particular the ongoing struggle for recognition from medical colleagues and the public, which forced them to actively define and defend their role:

> *“…the technical mindset allows us to be highly focused on diagnostic precision, but at the same time, we do not lose empathy for the patient. Still, I’ve been told by medical colleagues: ‘You can’t sign the reports you produce’ or ‘You can’t be called Doctor.’ But I was hired for my degree, my skills, and my right to perform this profession” (Ed.09.6);*
>
> *“…when I started, I didn’t even know who a radiology technician was. The general idea was just ‘the person who does X-rays in a dark room.’ Back then, and even now to some extent, the profession lacks a fully recognized identity” (Ed.010.4)*.
>
> *“…when I tell people what I study, they don’t understand what it is, I have to explain that it’s not nursing, not medicine, but however an healthcare professional. And that’s hard” (Stud.09.5)*.

Finally, through direct clinical practice, participants experience overcoming adverse situations and high-stakes clinical challenges as instrument of awareness. These defining moments linked technical vigilance with profound ethical responsibility:

> *“I was treating a patient, during a dental hygiene procedure, who wasn’t aware of having any systemic condition. Just by observing symptoms in the oral cavity, I was able to raise a red flag that led to the early diagnosis of an oral cancer” (Ed.17.6);*
>
> *“…it was a routine thyroid scintigraphy scan. My colleague noticed a barely visible anomaly on the image. We examined it further and told the physician, it turned out the patient had widespread papillary thyroid cancer. If she hadn’t raised her hand, that person would have died. That day, I understood what it means to walk on thin ice in healthcare, you can’t be afraid to fall in, because if you hesitate, someone else will drown” (Ed.014.6)*.

### 2.2. Element identified as crucial for professional identity

Participants identified two core elements that define the professional identity of a HTs: their hybrid nature and their capacity for continuous evolution and adaptability.

First and foremost, participants from both groups (n=33, 60%) strongly emphasized the **hybrid nature of the HT role**, which integrates technical duties with relational and assistance-focused responsibilities. They consistently positioned their profession beyond that of a mere operational technician. Students vividly described this dual identity in practice:

> *“…HTs are the ones who explain to the patient what is going to happen, who make them feel safe while working with machines. That’s what makes our role unique” (Stud.06.4);*
>
> *“…we’re not just technicians pressing buttons. We are there to support the patient, make them feel at ease, and still do a technically precise job” (Stud.09.2)*.

Educators elaborated on this, describing it as a “multifaceted identity” and coining the term “transversality” to capture their unique position as a bridge between technology and human care:

> *“…It’s a professional figure that interfaces with users, often unprepared and unaware of the procedures they are about to undergo, frequently frightened by the equipment. The most challenging but also the most stimulating part is this relational aspect. Then there’s the continuous need for updating, because as science and technology progress, we must keep up and master, or at least understand, those advancements to interact effectively with users and other health professionals. It’s a multifaceted identity, given how many aspects we are involved in” (Ed.03.5);*
>
> *“the radiology technologist is a figure that starts from a transversal base. Unlike other professions that are either technical or assistance-oriented, we are both. This transversality allows us to act as a bridge between medical figures and other healthcare professionals when working in teams. Many colleagues feel disappointed or demotivated and see this as a marginal profession. I see it as central to healthcare. For me, the identity of the radiology technologist is a core profession in healthcare, technically competent, relationally skilled, and versatile across disciplines. In one word: transversality” (Ed.005.4)*.

Second, participants identified **continuous evolution and adaptability** as a foundational component of their identity (n=19, 34%). They described their professional self as dynamic, not static, constantly being redefined by technological innovation and changing practice contexts:

> *“…The identity of the neurophysiopathology technician is a figure in strong growth… many new rehabilitative techniques have emerged… growth and change” (Ed.15.4);*
>
> *“…the identity of the radiology technologist is evolving… our role is highly influenced by the hospital we work in and the technologies we use…”* (Ed.06.5);
>
> *“..the identity of the audiometrist is very rich because the role is multifaceted: beyond performing diagnostic tests, we also engage in preventive actions, helping patients become aware of hearing risks. It’s a multifaceted identity. (Ed.09.4);*
>
> *“the perfusionist is a multidisciplinary profession. It spans from prenatal diagnostics to end-of-life care, including cardiac failure, ECMO, heart transplants, and organ donation. It’s a highly versatile figure” (Ed.07.4)*.

This evolution was also linked to a growing sense of professional autonomy over time. A senior educator reflected on his 35-year career, *“…the profession has grown from being an extension of the radiologist… to someone with autonomy, able to assert their role…” (Ed.02.4)*.

The dynamic and flexible nature was seen as the very essence of their identity, as one educator summarized, *“…our professional identity lies in a field that seems highly technical but is actually very dynamic and flexible… We are dynamic professionals with multiple skills” (Ed.14.4)*.

### 2.3. Professional Values

The construction of HTs professional identity was deeply rooted in core ethical values that shaped participants’ self-perception and clinical conduct.

**Respect for the human being** emerged as a foundational value guiding every interaction. More than two-thirds of participants (n=37, 67%) described this not as an abstract principle but as an embodied, relational ethic that required empathy and a conscious recognition of the patient’s vulnerability:

> *“…certainly, as a value, respect. And it can be summed up like this, when you work in close contact with a patient, you must be empathetic, enough to protect yourself too, and be able to manage the emotions tied to suffering, which otherwise would prevent you from performing and caring for the other. But I would summarize it with this: every time you meet a person, a man, a woman, a child, you should think of them as your brother, your sister, your parent, and respect what they’re facing in illness” (Ed.05.8);*
>
> *“…I mean I treat everybody the way I would like them to treat me. I know if it was my mother or father… who was a patient… that is the way I would treat my patients” (Stud.09.03)*.

This value was particularly crucial in high-tech or routine-driven environments where the risk of depersonalization was high, *“…we often act on autopilot and don’t realize we’re still dealing with a person. When the patient is asleep, it’s as if we forget they’re there… That is truly inappropriate” (Ed.15.8)*.

A smaller but sill important fraction of participants (n=18, 32%) emphasized the imperative to **ensure the quality of professional conduct.** This value extended beyond mere technical precision to encompass a profound sense of ethical obligation, accountability, and a commitment to “the look for truth” on behalf of the patient, *“…I suppose the look for truth, from an integrity perspective you’re looking… You have a person that has a question that needs to be answered” (Ed.05.45)*.

This commitment to quality was seen as especially critical amidst systemic pressures like staff shortages, which could lead to diagnostic exams, *“…then important values are precision and quality, because unfortunately, in this moment of crisis and shortage of healthcare staff, diagnostic exams are being performed poorly” (Ed.02.8)*. This sense of responsibility was deeply felt even in settings where there was no direct patient contact, as a laboratory-based professional explained, *“…behind the slides, behind the test tubes, behind the samples, there is a patient waiting for you to work seriously… so they can receive a diagnosis as soon as possible and begin treatment” (Ed.07.7)*.

Ultimately, this dual value system of respect and quality was described as extending beyond individual patients to encompass a broader responsibility to the healthcare team and the profession itself, *“…we are taking care about the patients and we are taking care about the professions and the professionals […] improving our technical fields” (Ed.07.03)*.

## 3. Construction of relational dimension of Educators and Students

The construction of the relational dimension emerged as a critical component of professional identity formation, describing this dimension not as a secondary skill but as a core practice that is learned, modeled, and negotiated within complex clinical and educational contexts.

### 3.1. Patient Interaction

The theme of **creating a comfortable environment** was voiced only by students (n=9, 60%), who identified it as a foundational element of care learned through observing their tutors. They described this as a prerequisite for patient trust and cooperation, involving emotional attunement, physical adaptation, and playful engagement in pediatric settings:

> *“…for example, by letting the child hold the instrument, they look at themselves in the mirror, make funny faces, play with me, and we involve the parent who is present… this really helps to make the child feel at ease and encourages cooperation” (Stud.02.10);*
>
> *“there are people with difficulties, and sometimes patients in wheelchairs couldn’t be placed in the chair, so in those cases, we had to adapt ourselves to the patient” (Stud.03.6);*
>
> *“…what struck me was the hygienist’s willingness to adapt to the patient, to welcome them. That really touched me and made me reflect. They really pay attention to the patient, especially if they have difficulties, and they show care through that attention” (Stud.03.7)*.

Second label emphasized by participants (n=26, 47%) is **integrating technical and relational care**, for example routine actions like explaining a procedure became acts of reassurance, *“I accompanied the patient to the diagnostic room, helped him lie down, and explained every action. That made it feel normal, it helped reassure him” (Stud.05.3),* or a conscious effort to resist depersonalizing professional language in daily practice, *“…sometimes I hear colleagues refer to patients by the exam they need, like ‘bring me the cranial CT’, but those are people on our scans. It’s something I’ve tried to correct in myself over time” (Ed.03.7) and “…the person in front of us isn’t just a diagnosis. If we refer them only as a case or a number, we fail to provide care” (Ed.02.08)*.

Students directly linked this relational connection to improved clinical performance, noting that it enhanced their confidence and technical execution, *“when I connect with the patient, the technical gesture goes better, I feel more confident doing it” (Stud.15.4)*.

**Adjusting communication to meet patient needs** was identified as an important skill for managing misinformation, dealing with diverse patient backgrounds, and explaining complex technologies.

> *“I adjust my communication based on the patient. It only takes a moment to understand. You need to be as clear and honest as possible, especially when there’s misinformation or fear, like common myths about radiation” (Ed.01.9)*.
>
> *“I work in the emergency department of a large city’s suburban area, and I often encounter patients from highly marginalized backgrounds who come to the ER for the most varied reasons. These are patients with a very low socio-economic status, and many of them have limited ability to speak Italian correctly and understand technical meanings” (Ed.01.10)*.
>
> *“Often patients arrive with very high expectations. I need to explain the process step by step, to lower those expectations and avoid disappointment. I say ‘this is the beginning of a journey we’ll do together’…” (Ed.22.11)*.

Students observed tutors modeling this “relational calibration” by adapting their tone and language to different patients*, “…with some patients, they used a more formal and clear tone, while with others they asked more personal questions, adjusting their voice, their tone, and the content of what they were saying. That helped patients feel more at ease” (Stud.01.6)*.

Finally, a theme voiced exclusively by educators (n=11, 20%) was the challenge of **striving to maintain a relational dimension of care** amidst systemic pressures. They described the constant tension between providing meaningful care and the constraints of time shortages.

> *“One thing I’ve always thought and tried to teach is that we need the patient’s collaboration to perform the exam as quickly as possible. That means welcoming the patient, gathering basic information, giving exam instructions, and addressing fears about radiation, in five minutes or less” (Ed.06.10)*.

Despite these limits, educators emphasized using time meaningfully, for instance by carefully explaining a consent form to a woman of childbearing age rather than just saying, *“sign here” (Ed.06.10)*.

### 3.2. Student Interaction

The way educators interact with students was identified as a core component of the educational process, shaping the professional identity of future HTs.

Educators emphasized the importance of **promoting student-centered teaching,** describing tailoring their methods to different learning styles and providing scaffolded support to guide students toward autonomy in the clinical setting, *“When these students arrive… at first I explain the work environment… Then, once they feel more confident, I suggest they try performing the exam independently. Of course, I follow them step by step until they reach autonomy… internships must be practical and grounded in real situations.” (Ed.02.11)*.

They understood that their student-centered teaching approach directly modeled the kind of professional-patient relationship they expected their students to develop, *“as educators, we are role models. And if we are not able to practice student-centred teaching ourselves, how can we expect to teach our students that the person should be at the centre?” (Ed.13.15)*.

They also spoke of intentionally **fostering positive attitude toward errors,** creating a safe relational climate where mistakes are not failures but as essential learning opportunities:

> *“…students shouldn’t be attacked when they make a mistake; they should be given the chance to fail in order to consolidate their learning. Mistakes are an integral part of training. In my opinion, if you never make mistakes, you never truly learn” (Ed.05.20)*.

## 4. Foundational elements of Educator role and Educational path

The data revealed several foundational elements that define the educator’s role and the ideal educational path for HTs, centered on pedagogical approaches that empower students and the institutional challenges that educators face.

### 4.1. Promoting Active Learning Strategies and Engaging Students

A cornerstone of the educational path was the promotion of active learning strategies that position students as protagonists in their own learning. Educators (n=29, 72%) described a necessary shift away from traditional, transmissive lectures toward more dynamic methods, *“…lecture cannot be just you like talk, talk, talk. But I think in this new age, it will be more interactive” (Ed.06.07)*.

To achieve this, educators suggest two main strategies:

1. Simulation and Case-based Learning: They used hands-on scenarios to replicate clinical practice, allowing *“them try play around with stuff, so if we do exposure factors for pain radiography just give them the phantom of the machine and if they ask how does it look with exposure, I said let’s try it out” (Ed.19.37),* in a safe environment *“so that […] students can make mistakes. And they can learn from mistakes but in a safe environment where mistakes cause no harm to patients” (Ed.15.9)*.
2. Gamification and virtual environments, *“…in the past, we didn’t have all the opportunities we have now […]. Today there is so much to learn in terms of didactic methodologies, the latest I’ve heard of is the use of an escape room using radiotherapy clinical cases to engage students. If I have to be honest, I feel I can use these methodologies, but I see them as something to be implemented only after a frontal teaching. Maybe I’m just old-fashioned” (Ed.02.18)*.

Underpinning these strategies was a philosophy of trust in the student’s agency, creating a climate of psychological safety where asking for help was encouraged and making mistakes was framed as an opportunity for growth, *“I think maybe sometimes I’m scared to make mistakes, even as a student. And you shouldn’t. You should be able to ask your colleagues for help” (Stud.07.08)* and *“…motivate them to do more next time when they come in, so when you engage with them and you’re like ‘do you want to try?’ […] they know that you’re going to cure the appreciation that they know what they’re doing” (Ed.23.22)*.

### 4.2. Promoting Hands-On Activities

A share of participants (n=17, 30%) underscored the critical need for hands-on activities to bridge the gap between theory and clinical reality. Educators saw this as essential for showing students how to perform tasks, a sentiment strongly echoed by students who called for *“more practical activities”* and *“small internships” (Ed.02.12; Stud.08.07; Stud.11.29)*.

However, educators identified significant barriers to providing these experiences, particularly the lack of financial investment for non-medical health professions, *“what is missing, are practical laboratories, or simulators. Very often, due to regulations, students are not allowed to operate manually in certain settings […] Unfortunately financial investment, is not considered for non-medical health profession degree programs” (Ed.05.23)*.

Despite these limitations, they highlighted the high value of even simple, well-structured laboratory exercises, *“I’ve seen students more involved when the explanation included videos or group-based practical exercises. Sometimes, I brought materials from my work lab to reproduce the preparation of drugs, syringes, vials, and coloured liquids for simulation” (Ed.13.12)*.

### 4.3. Promoting Critical-Reflective Thinking

Some participants (n=21, 38%) suggest the promotion of critical-reflective thinking for developing adaptive expertise. The goal was to help students move beyond procedural automatism to a state of internalized competence, where routine tasks become second nature, allowing for a greater focus on the patient, *“like riding the bike, you don’t think about it anymore, […] where a lot of it is automated and then you can put more focus on the patient” (Ed.16.3)*.

The path to this expertise, however, required conscious and deliberate reflection. Educators saw it as their role to nurture this mindset by explicitly aiming to *“develop students’ critical thinking skills” (Ed.16.19)* and by consistently asking the key reflective question *“What we can do differently?” (Ed.26.22)*.

### 4.4. Challenges in the Institutionalization of the Educator Role

Finally, educators described significant structural and institutional challenges, which they navigated with a strong sense of professional commitment and a desire for growth. Three main challenges emerged:

1. lack of financial recognition: a recurrent issue (n=20, 50%) was that teaching activities were often performed voluntarily, *“One thing that definitely needs improvement for healthcare technologists who teach on contract is the compensation. Often there is none, or it’s so small that it barely covers expenses” (Ed.09.16)*.
2. barriers to academic career: some educators (n=9, 22%) expressed a desire to pursue a PhD or a research career but encountered a system “that left them out”, *“I wanted to pursue an academic career after my Master’s degree, doing a PhD and becoming a researcher. At the time, there weren’t the opportunities and teaching in the university allows me, in a different way, remain in the academic and research environment” (Ed.20.09)*.
3. lack of formal pedagogical training: majority (n=26, 65%) reported entering teaching without structured preparation and expressed a clear desire to improve, *“It would help me a lot to have the chance to engage with someone more experienced in education and teaching, because I don’t know enough about these new methodologies. That’s why I want to keep learning as a teacher” (Ed.02.19).* Despite these barriers, educators demonstrated creativity and a proactive commitment to their own professional development, autonomously seeking out new teaching techniques *“…for me, updating is essential, especially on communication techniques and lesson delivery. Over the years, I’ve transformed my teaching, moving from traditional lectures to more interactive formats. I’ve learned from my experience with the Red Cross, using methods like role playing and gaming, especially when training non-healthcare participants.” (Ed.07.18)*.

## Discussion and future implication

This study reveals that the professional identity of HTs is a complex, dynamic, and socially constructed process. The findings illuminate the foundational role of motivation, the mechanisms of identity construction through practice, the ethical and relational core of the profession, the sophisticated pedagogical vision of its educators, and the significant institutional challenges that must be addressed.

The narratives of both students and educators reveal that career motivation is a foundational and multidimensional process in the formation of an HT’s professional identity. This process can be effectively understood through the lens of Self-Determination Theory (16), which distinguishes between intrinsic motivations (e.g., personal interest, a desire to help) and extrinsic motivations (e.g., influence of role models, social recognition). The participants’ testimonies provide rich examples of both, from students re-evaluating careers to find more “meaningful” work to those inspired by the passion of a family friend.

However, motivation in this context does more than simply propel an individual; it facilitates their entry into a professional community. This is where Social Identity Theory (17,19) becomes crucial, highlighting that professional identity emerges not just from personal psychology but from the internalization of a group’s shared values. The findings suggest that these diverse motivational pathways converge upon a central, shared value: the appreciation for the dual nature of the HT role. This “dual allegiance” to both scientific excellence and relational ethics becomes the defining trait of the HT community, allowing individuals to transform their unique biographical circumstances into a coherent professional self that is both personally fulfilled and socially embedded.

This study powerfully affirms that for HTs, professional identity is not a static attribute but a dynamic process of “building and rebuilding” (21), constructed through lived experience. The participants’ narratives show that clinical practice serves as the primary matrix for this formation, a concept that aligns strongly with Schön’s model of experiential learning and Lave and Wenger’s situated learning theory, where “doing is inextricably linked to becoming” (23).

This process is driven by a triadic articulation of doing, reflecting, and relating. Through *doing*, HTs confront clinical complexity and high-stakes challenges, which forge a resilient and ethically grounded sense of self (2). Through *reflecting*, they transform these lived experiences into professional knowledge, a process often catalysed by external feedback from peers, mentors, and students (8,24). Finally, through *relating*, HTs navigate their place within the healthcare ecosystem. This includes not only positive interactions, such as receiving validation from colleagues, but also the struggle against social misperceptions. This external friction, rather than hindering development, can paradoxically spur a deeper engagement with one’s role, compelling HTs to actively define their professional narrative (25).

The core elements of HT professional identity, its hybrid nature, adaptability, and vulnerability, are shaped by a dynamic interplay between internal self-perception and external social forces. The findings reveal a central tension: while participants define their identity by the unique duality of technical precision and human care, this “transversality” is often met with a lack of recognition (28). This discrepancy forces HTs into a constant process of identity negotiation and boundary work (39, 40). This aligns with social constructionist perspectives, where identity is a negotiated product of social structures (37), deeply embedded in situated contexts (36). As HTs navigate this fluid environment, they engage in a form of transformative learning (34), moving from passive executors to autonomous agents with a stronger sense of professional self-efficacy (38). Rather than retreating into rigid professional boundaries, many develop a “connective professionalism,” where legitimacy is built on collaboration and interdisciplinary teamwork (41).

The participants’ emphasis on respect as an embodied, active practice strongly resonates with philosophical conceptions of care, particularly Mortari’s (42) definition of attentiveness and Noddings’ (43) ethics of care. The “golden rule” ethos articulated by students demonstrates how moral identity is formed through the internalization of relational models that humanize both the caregiver and the person being cared for (44, 45). This study therefore argues that for HTs, relational ethics are not an adjunct to technical training but an essential humanizing force (46, 48).

This relational competence is multifaceted. It is learned and transmitted through a hidden curriculum (58), where students observe and replicate the tacit relational gestures of their tutors. This integration of relational and technical care directly impacts clinical efficacy, a finding supported by literature on emotional intelligence and adaptive performance (61). Central to this is adaptive communication, a dynamic clinical skill essential for procedural accuracy, patient trust, and co-constructing a therapeutic path (63,64,65). However, educators revealed a significant tension between this ideal and the “functional time” constraints of clinical practice (66), highlighting a nuanced professionalism where care is about using limited time meaningfully.

The dynamic construction of professional identity also involves the cultivation of aspirations for future development. Both students and educators articulated a vision of continuous evolution, grounding their sense of self in anticipated growth (20,50,51). Such future-oriented narratives reinforce the idea that professional identity is shaped through a transformative process of engagement with both personal motivation and institutional opportunity. The act of imagining one’s professional future functions as a structuring force in the present, guiding decisions, learning pathways, and relational attitudes. Supporting these trajectories is a critical educational responsibility to foster resilient, ethically anchored, and future-ready healthcare professionals.

The educators in this study reveal a sophisticated pedagogical vision aimed at shaping competent and reflective HTs. They are consciously moving towards a constructivist paradigm (79), using experiential learning (73), deliberate practice (74,75), and other active strategies to foster deep learning and psychological safety. These pedagogical efforts converge on a single, crucial goal: fostering critical-reflective thinking to move students beyond procedural automatism towards internalized competence and person-centered care (82,83,84).

However, this dedication to pedagogical excellence stands in stark contrast to the significant institutional challenges educators face, including a lack of financial recognition and limited pathways for academic advancement. Their perseverance fills a critical gap but is not a sustainable solution. Therefore, our findings point to clear and pressing implications for the future.

A key starting point is the development of national guidelines that formally recognize the distinct, hybrid role of HTs within Health Professions Education. Alongside this, there is a pressing need to invest in the pedagogical and relational preparation of HT educators, equipping them with the tools to foster the complex competencies identified in this study. Finally, to truly advance the status and quality of HT education, stronger international collaboration is needed to harmonize standards and encourage collective innovation. By addressing these areas, we can ensure that HTs are not only technically competent but also ethically empowered, relationally skilled, and fully recognized as vital contributors to the health professions.

## Conclusion

This study set out to explore the professional identity formation of Healthcare Technologists (HTs) within the Italian educational and clinical context, addressing a critical gap in the Health Professions Education (HPE) literature. HTs occupy a complex and often overlooked position in contemporary healthcare systems, straddling the domains of advanced technological expertise and direct patient care. Despite their centrality to clinical operations across a wide array of diagnostic and therapeutic contexts, HTs have historically remained peripheral to theoretical and empirical discussions surrounding professional identity formation, which have traditionally focused on physicians and nurses.

By conducting 55 in-depth interviews with both educators and students across multiple HT professional profiles, this study gives voice to a category of professionals whose perspectives are rarely cantered. The decision to focus on the lived experiences and self-reflections of HTs has allowed for the emergence of a nuanced and multilayered understanding of their identity development, rooted in everyday clinical practice, interpersonal dynamics, and the broader sociocultural frameworks in which they operate. This approach affirms that the construction of professional identity among HTs is not reducible to technical skill acquisition, but rather emerges through a continuous process of doing, reflecting, and relating, with patients, with peers, and with the profession itself.

Their testimonies reveal a hybrid identity that is both technically rigorous and ethically grounded, characterized by a desire to serve as mediators between increasingly complex medical technologies and the human experience of illness and care. In doing so, HTs not only contribute to diagnostic and therapeutic accuracy, but also play a critical role in supporting patients’ emotional well-being, fostering trust, and facilitating communication within multidisciplinary teams.

The voices of HT educators further highlight the challenges and potentials of teaching in this field. Many educators enter their roles with rich clinical backgrounds but limited pedagogical training, and yet demonstrate a strong commitment to student development, often acting as role models of compassionate, reflective, and adaptive professionalism. Their narratives also expose systemic issues, such as the lack of recognition, institutional support, and financial compensation, which must be addressed to fully institutionalize the educational mission of HTs and ensure the sustainability of quality teaching in this area.

Crucially, this study also sheds light on the experience of HT students, who navigate a training environment where they must learn to perform under pressure, communicate effectively, and manage emotional complexity, all while striving to define and assert their professional role within a healthcare system that does not always fully recognize or understand it. Their reflections reveal the importance of emotionally salient clinical experiences, of being seen and supported by educators, and of having the space to develop a coherent narrative about who they are becoming as healthcare professionals. The relational dimension of their training, including how they are treated by educators, how they interact with patients, and how they are perceived by other healthcare professionals, plays a central role in shaping their emerging identity.

In conclusion, the added value of this research lies in having made visible the invisible, by collecting and analysing the voices of those who are often left unheard. By bringing HTs to the centre of the conversation, we contribute to a more comprehensive understanding of what it means to become a healthcare professional today, in a world increasingly shaped by technological mediation and complex interpersonal dynamics. Future research and policy should build on these insights to ensure that HTs are not only technically competent, but also ethically empowered, relationally skilled, and pedagogically supported, fully recognized as vital contributors to the health professions.

## Data Availability

All data produced in the present study are available upon reasonable request to the authors

## Footnotes

2 Italian Ministry of Education, University and Research. Ministerial Decree no. 270 of October 22, 2004: Amendments to the regulations concerning the educational autonomy of universities. Official Gazette of the Italian Republic. 2004 Oct 23; General Series No. 245. https://www.gazzettaufficiale.it/eli/id/2004/11/12/004G0303/sg

3 https://www.tsrm-pstrp.org/index.php/codici-deontologici/

4 https://www.tsrm-pstrp.org/index.php/costituzione-etica-fno-tsrm-e-pstrp/

5 A detailed description of the governing regulations (Law 240/2010 and State-Regions Agreement 2007) is available in the supplementary materials.

6 **Coding Key:** *“Ed”* indicates that the speaker is an educator, while “Stud.” indicated a student speaker. The first number refers to the individual participant identification code, while the second number denotes the specific meaning unit within their interview transcript.

## Notes

### Competing Interest Statement

The authors have declared no competing interest.

### Funding Statement

This study did not receive any funding

### Author Declarations

Ethics committee of Human Sciences Department of University of Verona (Italy) gave ethical approval for this work

