## Supplemental Table 1 for "“Professional Identity at the Crossroads: Italian Healthcare Technologists navigating Technology and Human Care”"

### Full Coding System

<table>
<tr><th>EDUCATORS' LABELS</th><th>EDUCATORS' CATEGORIES</th><th>MACRO-CATEGORIES</th><th>STUDENTS' CATEGORIES</th><th>STUDENTS’ LABELS</th></tr>
<tr><td>Interesting in medical/healthcare field</td><td rowspan="4">CAREER MOTIVATION</td><td rowspan="9">PROFESSIONAL BACKGROUND</td><td rowspan="4">CAREER MOTIVATION</td><td>Interesting in scientific/medical field (anatomy, physiology, physics…)</td></tr>
<tr><td>Interesting in mixing technological and human aspect</td><td>Interesting in mixing technological and human aspect</td></tr>
<tr><td rowspan="2"></td><td>Changing from previous pathway</td></tr>
<tr><td>Knowing personally a HTs</td></tr>
<tr><td>Presenting professional role</td><td rowspan="3">PROFESSIONAL COURSE STORY</td><td>DESCRIPTION OF THE ACADEMIC JOURNEY</td><td>Presenting actual academic pathway</td></tr>
<tr><td>Reconstructing of past accademic path</td><td rowspan="4" colspan="2"></td></tr>
<tr><td>Structuring the educational path according to local policies</td></tr>
<tr><td>Motivation to pursue further studies</td><td rowspan="2">EXPERIENCES WITH POSTGRADUATE EDUCATION</td></tr>
<tr><td>Processing patway and challenges</td></tr>
</table>

| | | | | |
|---|---|---|---|---|
| Achieving through professional practice | **AWARENESS OF PROFESSIONAL IDENTITY** | **PROFESSIONAL IDENTITY BUILDING** | **AWARENESS OF PROFESSIONAL IDENTITY** | Achieving through professional practice |
| Achieving through educational pathway | | | | |
| Being a prof. figure with both technical and assistance features | **ELEMENT IDENTIFIED AS CRUCIAL FOR PROFESSIONAL IDENTITY** | | **ELEMENT IDENTIFIED AS CRUCIAL FOR PROFESSIONAL IDENTITY** | Being a prof. figure with both technical and assistance features |
| Being able to adapt and continuously evolve | | | | Being essential in the diagnostic and therapeutic process |
| Having multidisciplinarity competences | | | | Being a prof. figure with bridge role between patient and medical team |
| Improving pedagogical knowledge | **PROFESSIONAL GROWTH GOALS** | | **FUTURE PERSPECTIVE** | Becoming an educator |
| Improving socio-emotional competences | | | | Pursuing Post graduate Education |
| Updating technical skills | | | | Improving technical skills |
| Improving researching skills | | | | |
| Respecting of human being | **PROFESSIONAL VALUES** | | **PROFESSIONAL VALUES** | Respecting of human being |
| Ensuring the quality of professional practice | | | | |
| Adjusting communication to meet patient’s needs | **PATIENT INTERACTION** | **CONSTRUCTION OF RELATIONAL DIMENSION** | **PATIENT INTERACTION** | Adjusting communication to meet patient’s needs |
| Integrating Technical and Relational Care | | | | Integrating Technical and Relational Care |

| | | | | |
|---|---|---|---|---|
| Striving to ensure a relational dimension of care | | | | Creating a confortable environnement |
| Fostering a positive attitude toward errors | **STUDENT INTERACTION** | | **HEALTHCARE TEAM INTERACTION** | Promoting knowledge of the professional profile |
| Promoting student-centered teaching | | | | |
| Promoting active learning strategies and engaging students | **REQUIREMENTS TO ACTING THE EDUCATOR ROLE** | **FOUNDATIONAL ELEMENTS OF THE TRAINING** | **EDUCATIONAL PATHWAY** | Promoting hands-on activities |
| Promoting critical-reflective thinking | | | | Promoting feedback and peer-tutoring |
| Promoting hands-on activities | | | | |
| Evaluating Students’ Competencies | | | | |
| Being able to facing time and financial commitments | **CHALLENGES IN THE INSTITUZIONALIZATION OF THE EDUCATOR ROLE** | | | |
| Struggling to Enter Academia | | | | |
| Not Valuing Role Awareness Acquisition | | | | |
| Lacking Support for Educator Training and Updating | | | | |
